# Autologous cardiac micrografts as support therapy to coronary artery bypass surgery

**DOI:** 10.1101/2021.03.05.21252995

**Authors:** Annu Nummi, Severi Mulari, Juhani A. Stewart, Sari Kivistö, Kari Teittinen, Tuomo Nieminen, Milla Lampinen, Tommi Pätilä, Harri Sintonen, Tatu Juvonen, Markku Kupari, Raili Suojaranta, Esko Kankuri, Ari Harjula, Antti Vento, AADC consortium

**Affiliations:** Heart and Lung Center, Helsinki University Hospital and University of Helsinki, Finland; Department of Radiology, HUS Medical Imaging Center and Helsinki University Hospital and University of Helsinki, Finland; Päijät-Häme Central Hospital, Department of Internal Medicine, Lahti, Finland; Faculty of Medicine, Department of Pharmacology, University of Helsinki, Finland; Pediatric Cardiac Surgery, Children’s Hospital, Helsinki University Hospital and University of Helsinki, Finland; Department of Public Health, University of Helsinki, Finland; Department of Anesthesiology and Intensive Care, Helsinki University Hospital and University of Helsinki, Finland

**Keywords:** autologous micrografts, heart failure, coronary artery bypass surgery, cell therapy, atrial appendage, epicardial cell delivery

## Abstract

**Objectives:** Cardio-regenerative cell therapies offer additional biologic support to coronary artery bypass surgery (CABG) for treating the myocardium suffering from or damaged by ischemia. This phase 1, open-label study assessed the safety and feasibility of epicardial transplantation of atrial appendage micrografts (AAMs) in patients undergoing CABG surgery.

**Methods:** Twelve consecutive patients destined for CABG surgery were included in the study. Six patients received AAMs during their operation and six patients were CABG-operated without AAMs treatment. Data from 30 elective CABG patients was collected for a conjunctive control group. The AAMs were processed during the operation from a biopsy collected from the right atrial appendage. They were delivered epicardially on the infarct scar site identified in preoperative CMR. The primary outcome measures at six-months follow-up were i) patient safety in terms of hemodynamic and cardiac function over time and ii) feasibility of therapy administration in a clinical setting. Secondary outcome measures were left ventricular wall thickness, change in myocardial scar tissue volume, changes in left ventricular ejection fraction, plasma concentrations of N-terminal pro-B-type natriuretic peptide (NT-proBNP) levels, NYHA class, number of days in hospital and changes in the quality of life.

**Results:** Epicardial transplantation of AAMs was safe and feasible to be performed in conjunction with CABG surgery. CMR demonstrated an increase in viable cardiac tissue at the infarct site in patients receiving AAMs treatment.

**Conclusion:** Transplantation of AAMs shows good clinical applicability as adjuvant therapy to cardiac surgery and can additionally serve as a delivery platform for cardiac gene therapies.

**Trial Registration:** ClinicalTrials.gov identifier NCT02672163

## 1. Introduction

Coronary artery bypass graft (CABG) surgery reinstates myocardial blood flow downstream of an occluded coronary artery. Although CABG provides the patient with symptomatic benefit, it unfortunately does not alone restore the cells lost to infarction.^1, 2^ Adjuvant or support therapies, including regenerative cell transplantation, have therefore been investigated for decades, but none have so far been adopted for clinical use.^3, 4, 5^ Furthermore, many therapies are associated with lengthy processing times and significant costs. ^6^

The current consensus for a CABG-supportive cardioregenerative therapy is that the treatment should consist of a mixture of cardiac cell types and their extracellular matrix in a synergistic composition.^7, 8, 9^ To this end, we have demonstrated the preclinical efficacy and initial clinical feasibility^10^ of autologous cardiac microtissue, atrial appendage micrografts (AAMs), and have shown that AAMs therapy activates cardiogenic and cardioprotective pathways. ^11^ Based on these encouraging results, we initiated a phase I open-label, non-randomized clinical study ^12^ to evaluate the safety and feasibility of AAMs transplantation in conjunction with CABG surgery. We utilized gadolinium-enhanced cardiac magnetic resonance (CMR) imaging before and six months after treatment to gain detailed insight into the AAMs ‘efficacy to induce myocardial repair.

## 2. Methods

### 2.1 Ethics and patient selection

The study protocol was evaluated and approved by the Surgical Ethics Committee of the Hospital District of Helsinki and Uusimaa (number 180/13/03/02/13). The study is registered in the ClinicalTrials.gov database with the identification number NCTNCT02672163. For the interventional phase I, open label non-randomized study, a total of 12 patients scheduled for elective CABG surgery from Helsinki University Hospital, Finland were recruited in chronological order. An additional 30 patients served as site- and time-matched reference controls. Similar to our earlier clinical cell therapy trial evaluating bone marrow mononuclear cell transplantation ^13, 14^, patients of either gender were evaluated for participation if they had ischemic heart failure and were scheduled for elective CABG. The criteria for inclusion and exclusion of the patients are presented in *Table 1*. Each patient was given both oral and written information about the trial, and the patient ‘s written informed consent was required for participation. After recruitment and drug optimization, patients waited four to twelve weeks for the elective operation. During this time, the ejection fraction (EF) changed in the AAMs group and control group.

**Table 1:**
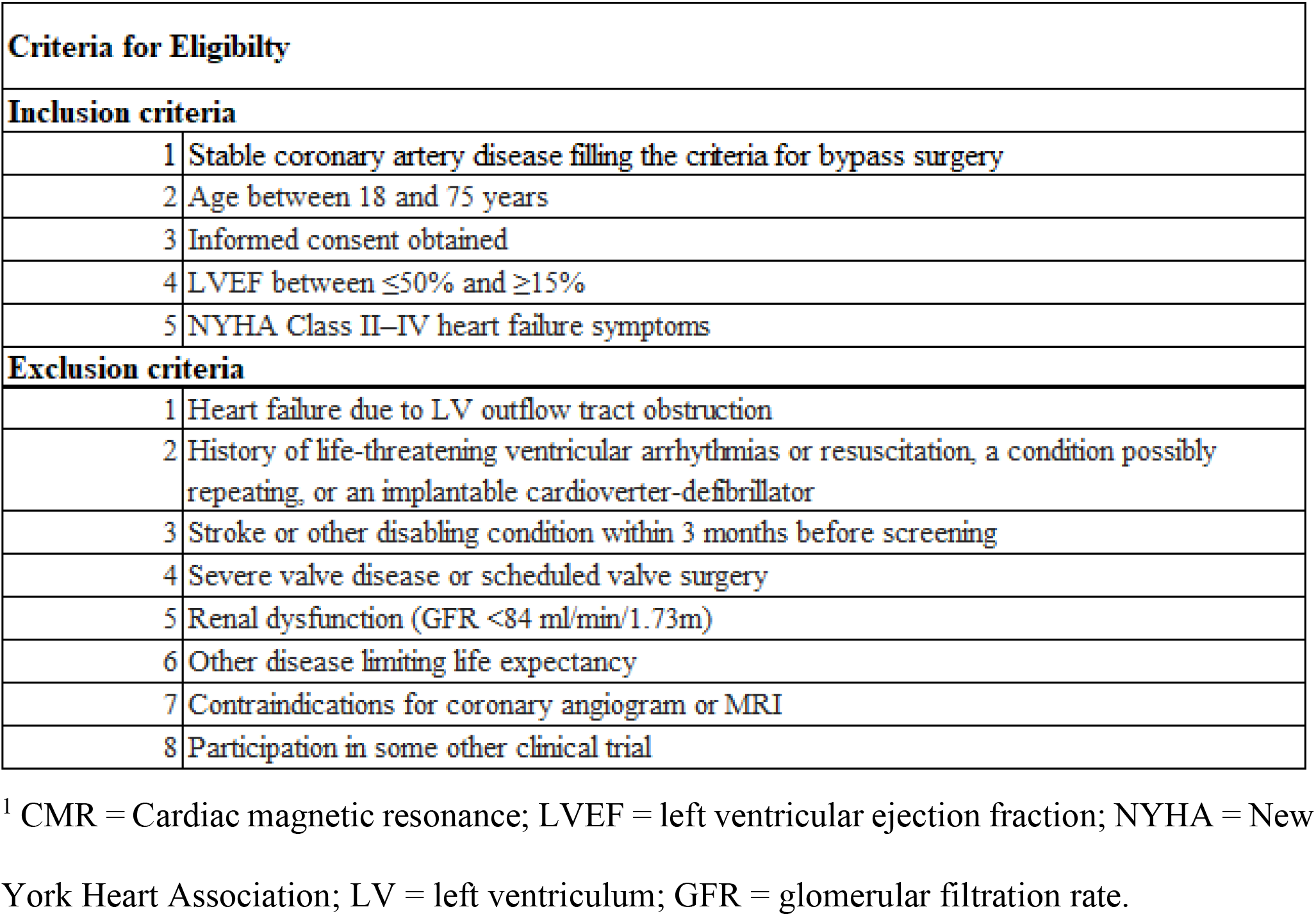
Criteria for patient selection. Inclusion and exclusion criteria for patients enrolled in the study.

The first six patients were recruited to the AAMs group and they received the AAMs transplant during CABG surgery. The next six patients formed Control group I. Their surgery was performed according to normal hospital protocol without the AAMs transplant. These two groups underwent the same follow-up protocol. To determine more on the safety of the procedure, the following thirty patients who met the study ‘s inclusion and exclusion criteria, and were scheduled for elective CABG operation, formed Control group II. These patients were treated according to normal hospital protocol, without the AAMs transplant and any additional imaging, examination or blood tests required for the prior two groups.

Echocardiography (echo), quality of life (QoL) as measured using the health-related questionnaire instrument 15D^© 15^, New York Heart Association (NYHA) class, basic laboratory tests, and blood N-terminal pro-B-type natriuretic peptide (NT-pro-BNP) were evaluated at baseline, as well as at three and six months follow-up for the patients in the AAMs group and Control group I. CMR was performed twice per patient: at the pre-trial and at six months follow-up. The study outline is shown in *Figure 1A*.

**Figure 1.**
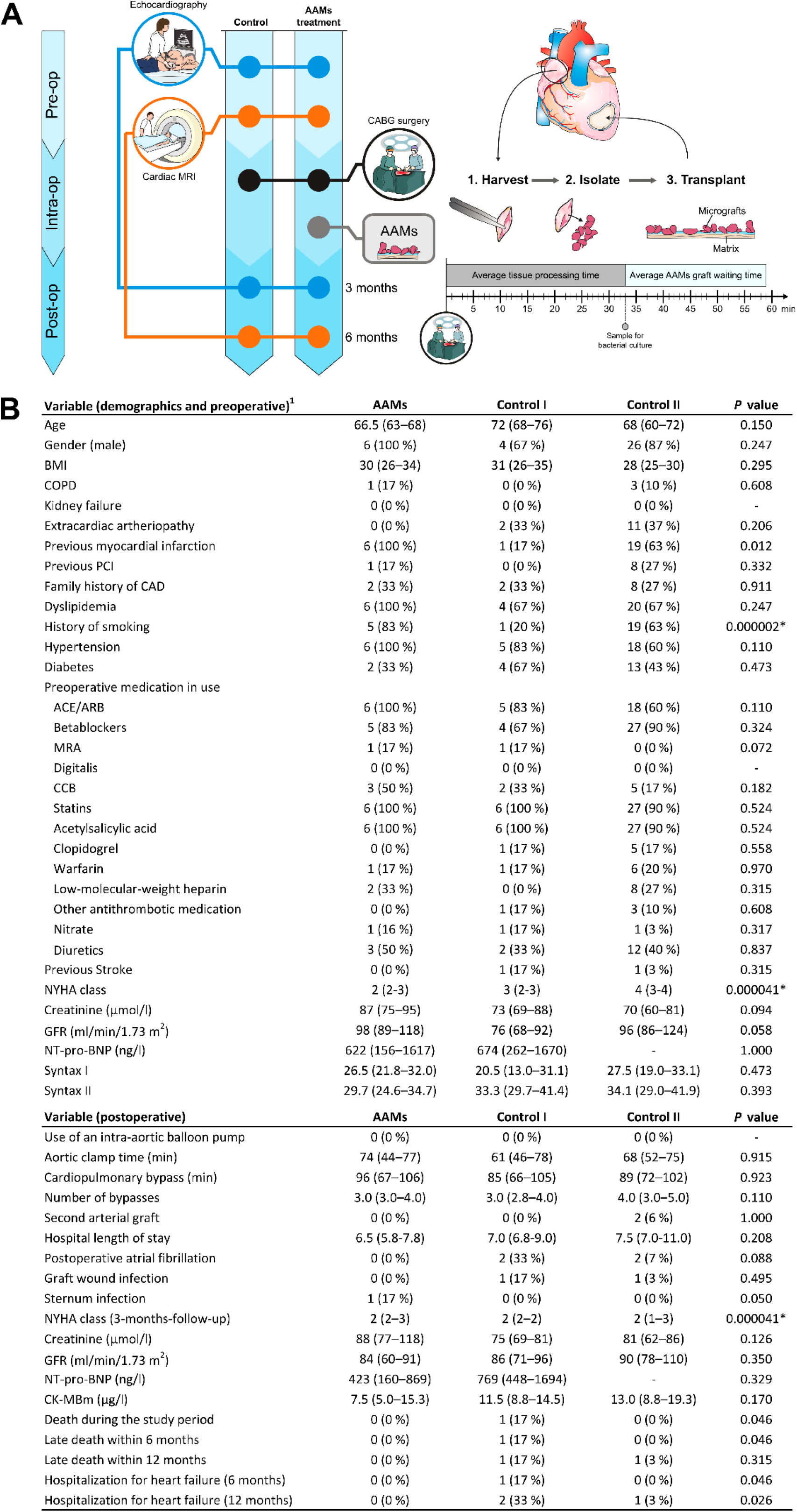
Study outline and Demographics. Study outline and demographics with pre- operative and post-operative data. *(A)* All patients underwent pre-operative (Pre-op) echocardiography and CMR. The Study group received AAMs therapy during coronary artery bypass (CABG) surgery, while the Control group I (Control I) only underwent CABG surgery. AAMs were harvested from the right atrial appendage, mechanically isolated, and transplanted intraoperatively (Intra-op). Patients in both groups were examined three-months post- operatively (Post-op) by echocardiography and six months after the operation using CMR. *(B)* Demographics, pre- and postoperative data of the AAMs group (AAMs) and both control groups (Control I and Control II). ^1^ ACE, angiotensin-converting enzyme inhibitor; ARB, angiotensin receptor blocker; BMI, body mass index; CAD, coronary artery disease; COPD, chronic obstructive pulmonary disease; CCB, calcium channel blockers; CK-MB, creatine kinase myocardial band; GFR, glomerular filtration rate; MRA, mineralocorticoid receptor antagonists; NT-Pro-BNP, N-terminal pro-B-type natriuretic peptide; NYHA, New York Heart Association; PCI, percutaneous coronary intervention. Results are presented as median (IQR), group comparisons were performed with the Kruskal-Wallis test. Mann Whitney U test was used for the variables in only Study and Control group I results, or Chi Square for ordinal variables. Significant results after correction (Bonferroni) for multiple comparison are marked by an asterisk (*), with the results of post-hoc between-group analyses found in the text.

### 2.2 Micrograft isolation

During CABG surgery, AAMs were harvested from the right atrial appendage. A small piece of atrial appendage was removed while inserting a venous cannula for the heart-and-lung machine. The size and quality of appendage differs between patients according to age, gender, and comorbidities. To standardize the removable size of atrial appendage tissue, a minimum length of 5 x 10 mm and weight from 600 mg to 800 mg was required for each sample.

The harvested tissue was processed on-site in the operating room using a cell therapy tissue homogenizer (Rigenera-system, HBW s.r.l., Turin, Italy). Cell isolation was performed by a nurse who had received training in a cell-culture laboratory and in our previous studies by using a large-animal model (manuscript in preparation, Nummi et al.). Isolation was performed under strict adherence to sterility. We have previously described the preparation of the AAM transplant in detail. ^10, 12^

The isolated AAMs were applied in cardioplegia suspension to the extracellular matrix sheet (Cormatrix**®** ECMTM Technology, Cormatrix Cardiovascular Inc., Atlanta, GA, USA). Fibrin sealant (Tisseel^TM^, Baxter Healthcare Corp. Westlake Village, CA, USA), routinely used in surgery for tissue glue, was added to the cell suspension to secure the AAMs to the matrix. The sheet with AAMs was placed on top of the damaged myocardium so that the cell surface was facing the epicardium. The Cormatrix® attached well to the tissue but was further secured to the myocardial surface by three to four simple sutures with nonabsorbable monofilament polypropylene string.

### 2.3 Therapy administration

A standard CABG operation was performed under cardiopulmonary bypass and mild hypothermia, where patients were under cardiac arrest and receiving cardioplegia protection. After completion of the bypass anastomoses, the AAMs transplant was placed over the infarct scar area as determined preoperatively from individual patient ‘s pre-CABG CMR images. The therapy application procedure was carefully photographed during each surgery, and the treatment administration site was meticulously detailed in patient documents for further CMR and echo analyses.

### 2.4 Tests for transplant sterility

To verify sterility, samples for microbial cultures were taken from each AAM-transplant.

### 2.5 Clinical Cardiac CMR

CMR imaging was performed with a 1.5 T Avanto fit scanner and phase array cardiac coil (Siemens, Erlangen, Germany). Images were electrocardiography gated and taken during breath-holding. LV structure and function were imaged by a standardized CMR protocol. TrueFISP cine series was obtained at the vertical and horizontal long axis for scouts to line up short-axis images. The stack of short-axis images was obtained from the mitral valve plane through the apex. To detect the myocardial scar, late gadolinium enhancement (LGE) was imaged with a 2D-segmented inversion recovery gradient echo sequence 12 to 20 minutes after Dotarem® injection (279.3 mg/ml; dose 0.2 mmol/kg). LGE images were obtained for the same views and slice/gap thickness as cine imaging.

All images were analysed using a dedicated workstation (Medis® suite 3.2.28.0. Medis Medical Imaging Systems, Leiden, The Netherlands). LV function and volumetry were assessed from cine images with Qmass Analyse Program analysis software (Medis) and global longitudinal strain with Qstrain (Medis). Full width with half maximum (FWHM) technique and signal threshold versus reference mean (STRM) threshold of 5 standard deviations (SD) above the mean signal intensity (SI) was used to assess myocardial and infarcted area mass. The thickness of infarction was measured from the thinnest point exactly on the same position from both images.

### 2.6 End-point measures

The primary outcome measures were patient safety in terms of haemodynamic and cardiac function and feasibility of the therapy administration in a clinical setting. Haemodynamics were evaluated during the operation and stayed at the intensive care unit (ICU) given requirement for vasoactive medication and success of weaning from cardiopulmonary bypass and respirator. Postoperative hemodynamic criteria for assessing safety were cardiac index, haemoglobin, central venous oxygenation (SvO_2_), serum potassium level, blood glucose and lactate levels, and arterial pH. Cardiac function was evaluated during the operation by transoesophageal echo and during the stay at ICU by transthoracic echo as well as constant telemetric monitoring of rhythm. Criteria for peri- and post-operative myocardial infarction was new regional wall motion abnormality confirmed by echo, ck-mb > 50 μg/L, ischemic changes in ECG (LBBB or Q-waves), ventricular arrhythmia, or angiographically documented new graft or new native coronary artery occlusion. Feasibility was evaluated by the success in completing the delivery of the cell sheet to the myocardium, waiting times in minutes for the AAMs transplant and the success in closing the right atrial appendage by purse-string suture without additional sutures or patching.

The secondary outcome measures were change in LV infarct area thickness, movement and diastolic function assessed by CMR, change in the amount of myocardial scar tissue as assessed by CMR, local changes in systolic and diastolic measures as estimated by echo, changes in LVEF, pro-BNP level, NYHA class, hospitalization or the days in hospital and QoL.

### 2.7 Statistics

Results are given as median with interquartile ranges (IQR), or *n* (%) of group for ordinal and nominal values. Comparisons between groups were performed with the Mann Whitney *U* test, or between three groups with the Kruskal-Wallis test. Ordinal variables were tested with the Chi Square test. Multiple comparisons were corrected with the Bonferroni method, significant findings were further tested groupwise using the Mann Whitney *U* test or Chi Square test, as applicable. Quality of life data is presented as mean (SD), and were analysed with the independent samples *t*-test (two-sided). The level of significance (α) was 0.05. Analyses were performed with the IBM SPSS Statistics 25 program (IBM Corp., Armonk, NY).

## 3. Results and discussion

Patient demographics are presented in *Figure 1B*. The median NYHA class differed between the AAMs group and Control group II (median NYHA 2 (range 2 to 3) in the AAMs group and median 4 (range 2 to 4) in the Control group II; *P* < 0.0001). Control group I had NYHA median 3 (range 2 to 3). A history of smoking was more common in the AAMs group (*N* = 5, 83%) than in the control groups (Control I, *N* = 1, 20%; Control II, *N* = 19, 63%; *P* < 0.0001). Otherwise, there were no significant differences between the groups.

All atrial appendages were closed without difficulties with a single purse-string suture. The AAMs sheet was easily prepared in the operating room during CABG surgery. The median time for preparing the AAMs sheet was 33 min (range 22 to 43 min), and the median waiting time for the sheet was 26 min (range 7 to 54 min). In every operation, the AAMs sheet was ready to be placed before the anastomosis was done. The sheet was secured with three to four sutures without difficulties. Bacterial cultures taken from each sheet were negative for bacterial growth, demonstrating the sterility of the procedure.

Physiological data from the intensive care unit (ICU) at admission and 24 h after are shown in *Table 2*. Despite a significant difference in 24 h lactate concentrations between the three groups, all lactate values were within normal range (0.8 to 1.1 mmol/L). Echocardiography measurements and ECG findings demonstrated no significant differences between the study groups (*Table 3*).

**Table 2:** Physiological data from ICU. Physiological data from the first 24 hours of the ICU, comparing the study group to the two control groups. No significant differences were noted between the three groups. Results presented as median (IQR), group comparison performed with the Kruskal-Wallis test and Bonferroni correction (corrected significance level *P* = 0.001).

| Variable | Time | AAMs | Control I | Control II | p |
| --- | --- | --- | --- | --- | --- |
| Heart Rate ($\text{min}^{-1}$) | 0 h | 53 (42–69) | 73 (61–82) | 85 (78–89) | 0.03 |
| CVP ($\text{cmH}_2\text{O}$) | | 8 (5–9) | 8 (4–11) | 10 (6–13) | 0.293 |
| Systolic arterial pressure (mmHg) |  | 112 (106–136) | 113 (96–129) | 105 (93–113) | 0.138 |
| MAP (mmHg) |  | 79 (71–87) | 74 (65–83) | 67 (63–77) | 0.128 |
| Diastolic arterial pressure (mmHg) |  | 61 (55–66) | 60 (47–62) | 55 (48–59) | 0.094 |
| SpO<sub>2</sub> (%) |  | 100 (99–100) | 100 (94–100) | 99 (98–100) | 0.368 |
| aB-BE |  | 0.1 (-1.0 – 0.8) | -2.2 (-3.0 – -0.6) | -0.7 (-2.1 – 0.8) | 0.166 |
| Lactate (mmol/l) |  | 1.3 (1.2–1.8) | 1.3 (0.9–1.6) | 1.0 (0.8–1.4) | 0.188 |
| Hb (g/l) |  | 136 (123–141) | 128 (93–128) | 115 (99–133) | 0.162 |
| Glucose (mmol/l) |  | 7.1 (6.4–9.4) | 9.0 (6.6–12) | 7.3 (6.6–10.3) | 0.681 |
| Heart Rate ($\text{min}^{-1}$) | 24 h | 82 (77–92) | 85 (78–95) | 82 (78–90) | 0.793 |
| CVP ($\text{cmH}_2\text{O}$) | | 10 (8–13) | 7 (5–10) | 10 (6–14) | 0.342 |
| Systolic arterial pressure (mmHg) |  | 143 (120–157) | 123 (118–137) | 114 (102–130) | 0.009 |
| MAP (mmHg) |  | 87 (77–88) | 75 (73–82) | 69 (66–81) | 0.043 |
| Diastolic arterial pressure (mmHg) |  | 61 (56–62) | 55 (51–61) | 52 (46–59) | 0.112 |
| SpO<sub>2</sub> (%) |  | 97 (96–99) | 99 (96–99) | 98 (96–99) | 0.751 |
| aB-BE |  | -1.0 (-1.7 – 0.6) | -1.6 (-2.3 – -0.25) | -1.5 (-3.2 – -0.2) | 0.451 |
| Lactate (mmol/l) |  | 1.0 (0.8–1.1) | 0.6 (0.6–1.0) | 1.2 (1.0–1.7) | 0.005 |
| Hb (g/l) |  | 99 (93–108) | 111 (92–120) | 107 (96–115) | 0.590 |
| Glucose (mmol/l) |  | 8.2 (7.7–8.3) | 8.7 (7.1–12.3) | 8.1 (7.0–8.9) | 0.651 |

<sup>1</sup> CVP = Central venous pressure; MAP = mean arterial pressure; SpO<sub>2</sub> = peripheral capillary oxygen saturation; aB-BE = arterial Astrup base excess; Hb = haemoglobin.

**Table 3:**
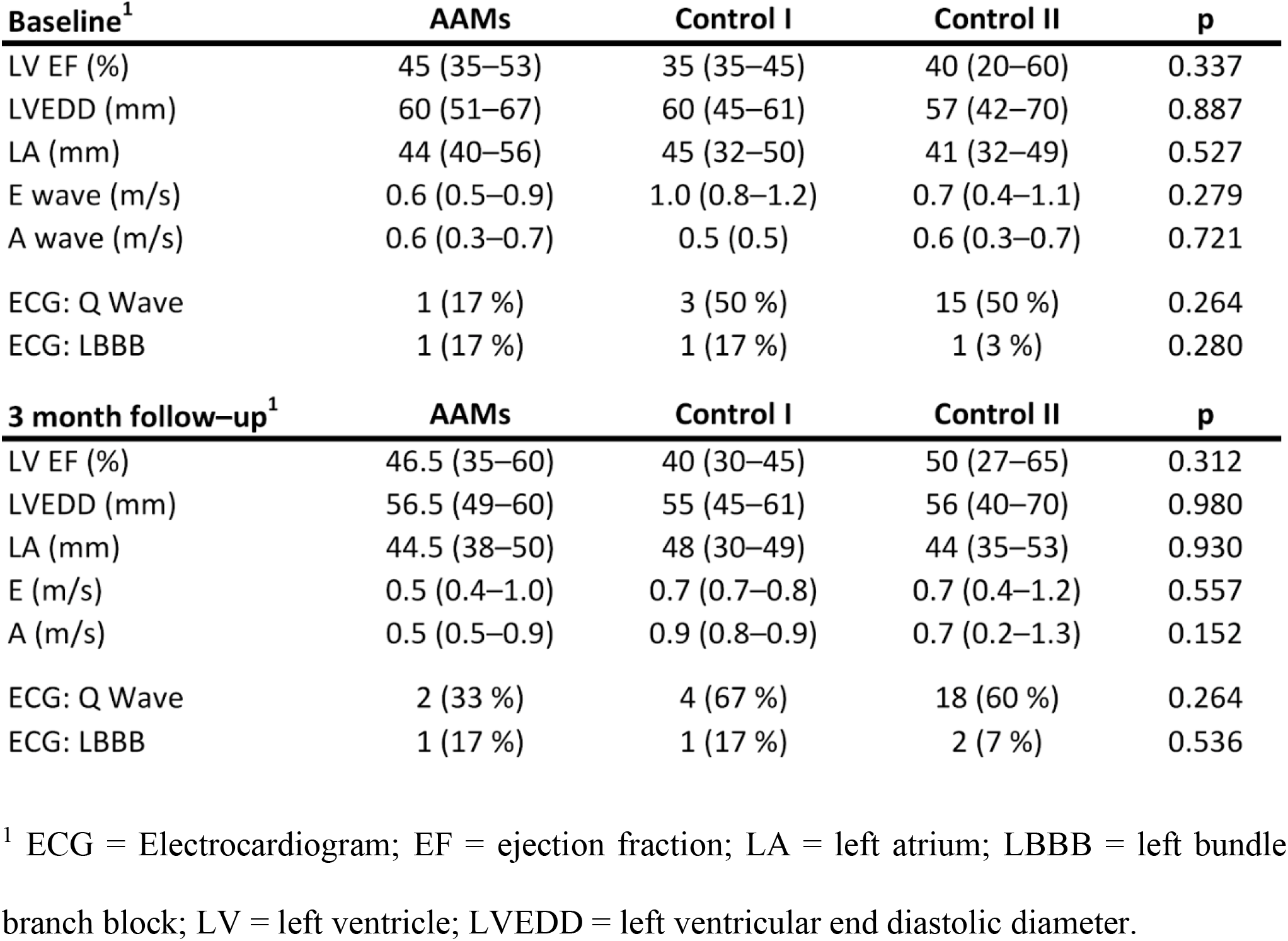
Cardiac ultrasound and electrocardiogram data. Cardiac ultrasound and electrocardiogram data from the preoperative baseline to the 3-month check-up visit, with comparisons of the study group versus both control groups. No significant differences were noted between groups. Results presented as median (range), group comparison performed with the Kruskal-Wallis test or Chi Square test (ordinal variables) with Bonferroni correction (corrected significance level p = 0.007).

**Table 4:**
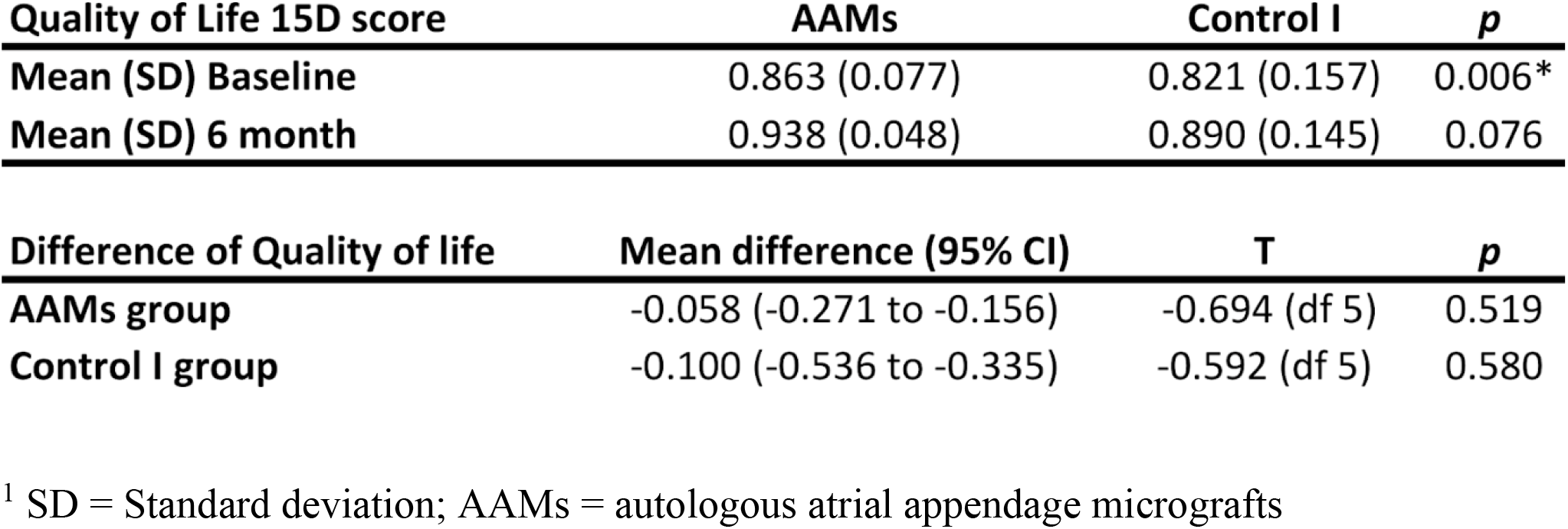
Quality of life comparison. Quality of life of the study and Control I group at baseline and at the 6-month follow up, with groupwise comparison. The mean difference comparison of both groups against the reference group of the age-matched healthy general population at baseline and at the 6-month follow-up is presented in the lower table. Results presented as mean (SD), mean difference is presented with 95% CI and groupwise comparisons were performed with the Student ‘s *t*-test.

Postoperative complications are presented in *Figure 1B*. One patient from the AAMs group was diagnosed with sternum-wound infection postoperatively. The patient recovered from the infection with antibiotics and no further operative procedures were required. There were no sternum-wound infections in the control groups. Graft wound infections were diagnosed from two patients in Control group I and one patient in Control group II. There were no strokes or myocardial infarctions during the study period in any of the three groups.

Discharge time from the hospital was similar between the groups. Readmission to hospital due to heart failure occurred more often in Control group I than in the AAMs group (N = 0, 0%; Control I N = 2, 33%; and Control II *N* = 1, 3%, *P* = 0.02). There were no deaths in the AAMs group during the study period of 6 months or the follow up time of one year. One patient from Control group I died of systolic heart failure during the study period (*P* = 0.046) and was lost to follow up.

CMR imaging (*Figure 2*) was performed on patients from the AAMs group and Control group I preoperatively and at 6-months follow-up. Infarct scar was detected in all patients ‘preoperative CMR analyses. Change of viable myocardial thickness at the infarcted area demonstrated a significant increase in the AAMs group (1.0 mm (range 0.2 to 1.3 mm) and Control group I (−1.4 mm (range −1.7 to 0.0 mm), *P* = 0.009). The mainly preferred method of evaluation is the STRM of 5SD gives better approximation of the extent of heart infarct as compared with FWHM ^16, 17^ but as previous studies often used the FWHM, these results are also presented in *Figure 2*.

**Figure 2.**
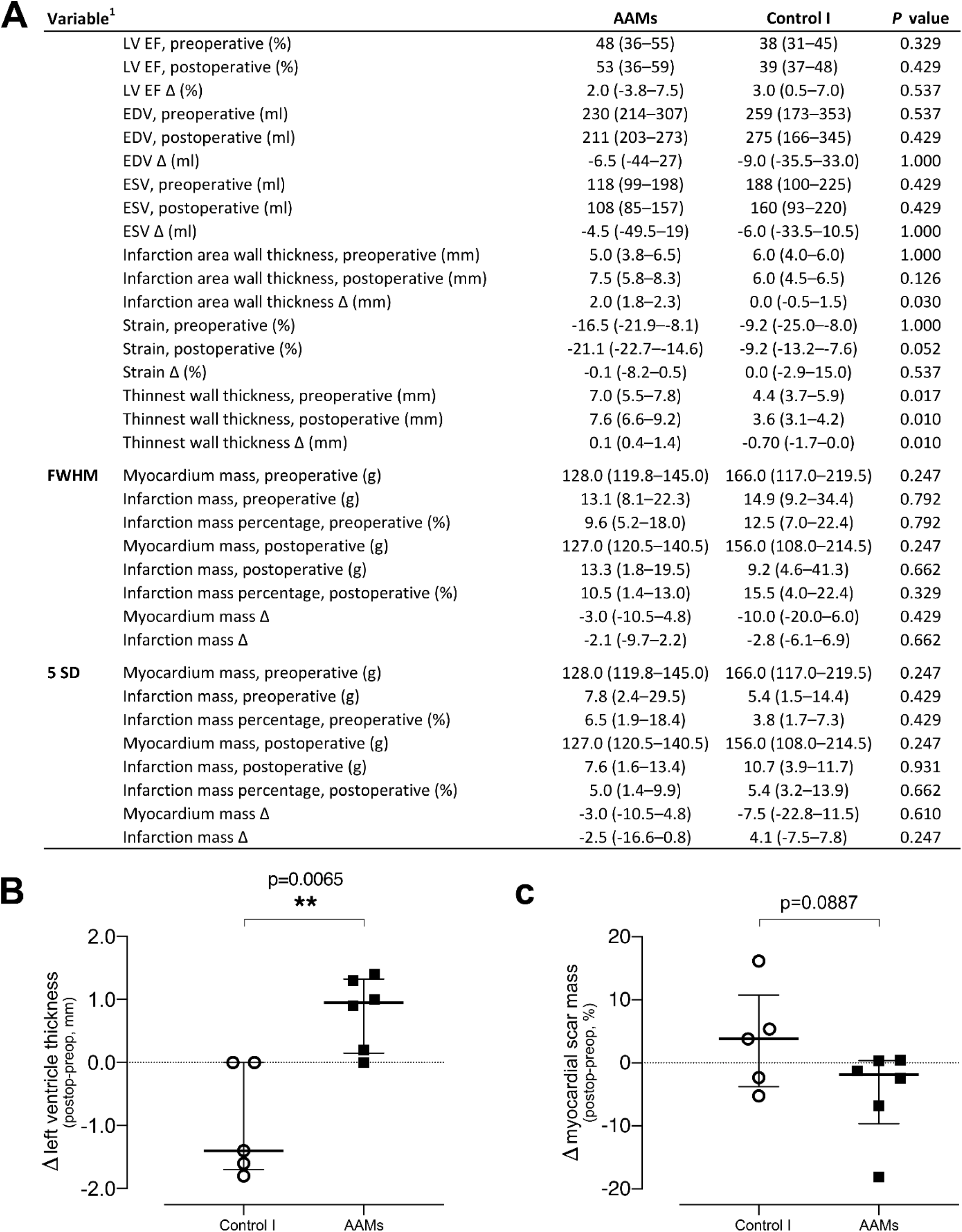
Cardiac magnetic resonance imaging. Evaluation of function and structure by cardiac magnetic resonance imaging. *(A)* Comparison of the AAMs group patients with the Control group I patients. For all variables, a preoperative and postoperative value is presented, followed by the absolute change (Δ) of these values. Results presented as median and IQR, group comparison performed with the Mann Whitney *U* test (Bonferroni adjusted *P* of 0.002). *(B)* Single parameter comparison of postoperative-preoperative change in viable left ventricle thickness at infarct scar site. *(C)* Single parameter comparison of postoperative-preoperative change in myocardial scar mass. ^1^EDV, end diastolic volume; EF, ejection fraction; ESV, end systolic volume; LV, left ventricle; FWHM, full width with half maximum; SD, standard deviation.

**Figure 3.**
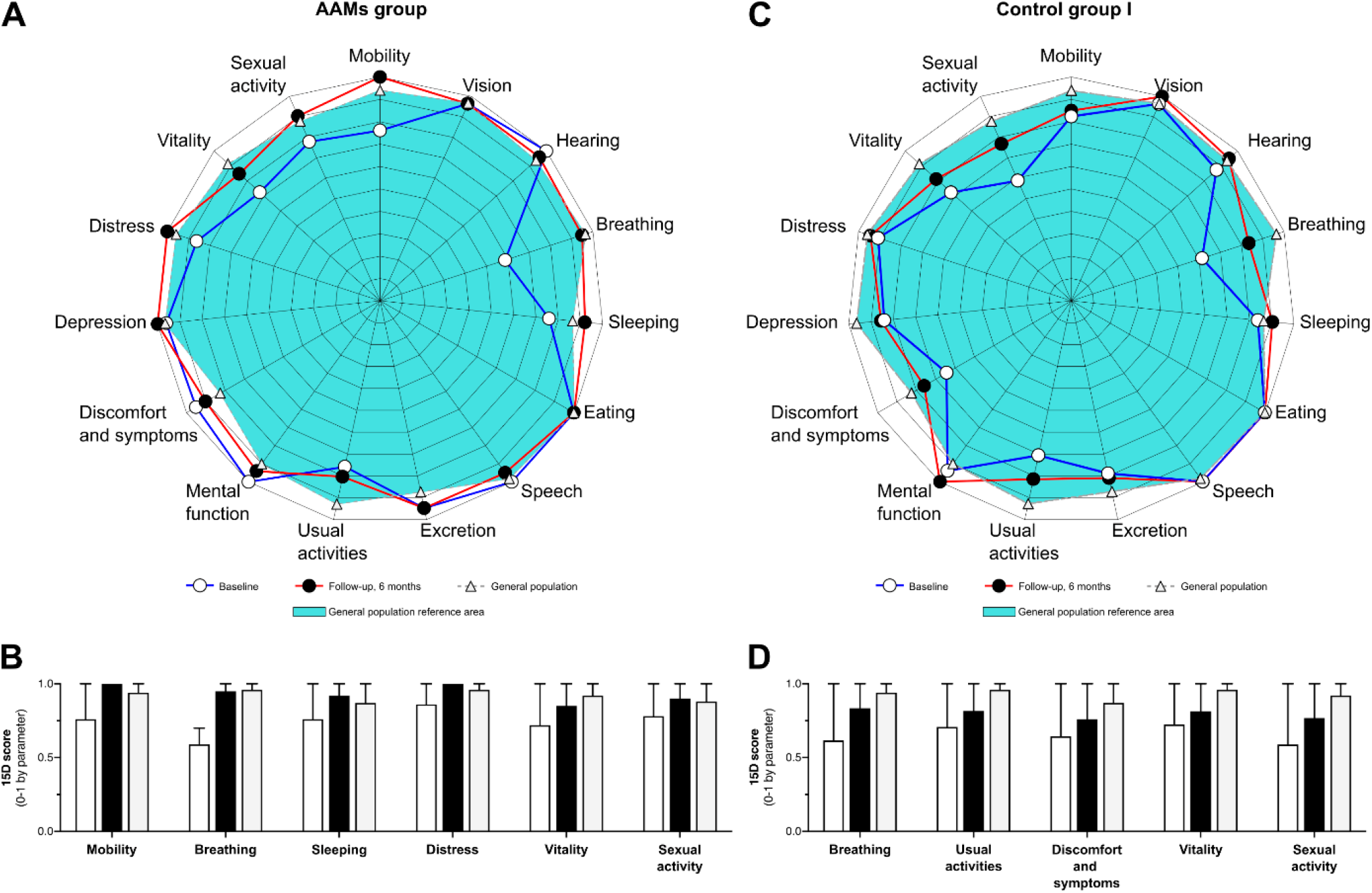
Gaphical presentation of quality of life. Health-related quality of life QoL in the AAMs group and the Control group I. *(A)* Graphical presentation of 15D QoL scores preoperatively and at the 6-month follow-up in the AAMs group. *(B)* Selected parameters of the 15D QoL in the AAMs group. White bars represent preoperative values, black bars represent values at 6-months follow-up and grey bars represent the respective parameter ‘s 15D QoL score in the general population. *(C)* Graphical presentation of 15D QoL scores preoperatively and at the 6-month follow-up in the Control group I (Control I). *(D)* Selected parameters of the 15D QoL in the Control I group. White bars represent preoperative values, black bars represent values at 6-months follow-up and grey bars represent the respective parameter ‘s 15D QoL score in the general population.

The Health-related QoL of the surviving AAMs and Control I patients did not differ significantly at baseline or at 6 months. Similarly, neither group ‘s QoL differed significantly from that of the reference population, that is, age-matched samples of the general population. The individual mean 15D scores for both groups are presented in *Table 3*. At the 6-month follow-up, the Study group 15D Index score mean (SD) of 0.8634 (0.07685) was slightly better than the Control I group 15D

Index score mean (SD) of 0.8207 (0.15693). The mean (95% CI) difference was 0.04267 (−0.11628 to 0.20162); independent t-test T(10) = 0.598, *P* = 0.006. According to the data of Sintonen et al. ^15^, the Finnish age-matched adult general population (n = 562) has a mean (SD) 15D HRQoL index score of 0.9210 (0.07611). Compared to the reference population, the study group (n = 6) had a mean (95% CI) difference of −0.06 (−0.27 to 0.16); independent t-test T(5) = −0.69, *P* = 0.519. The Control I group (n = 5) mean (95% CI) 15D index score differed −0.10 (−0.54 to 0.34); independent t-test T(5) = −0.59, *P* = 0.580.

We conclude that epicardial transplantation of AAMs is safe and feasible to be performed in conjunction with CABG surgery in the operating room. These results warrant further evaluation of the AAMs treatment ‘s efficacy in a larger randomized controlled trial with changes in myocardial function and structure by gadolinium-enhanced CMR imaging as the primary efficacy endpoint. The combination of intraoperative harvesting, isolation and transplantation of AAM cardiac tissue shows promise as a safe, straightforward, and clinically feasible support therapy to CABG surgery.

### Clinical perspectives

In this non-randomized, open label trial epicardial delivery of AAMs were safe and feasible in conjunction of CABG surgery. Cardiac magnetic resonance showed thickening of the left ventricular wall in site of infarct compared with CABG surgery alone. Transplantation of AAMs shows good clinical applicability as adjuvant therapy to cardiac surgery, and may serve as a potential delivery platform for future cell or gene-based cardiac therapies.

### Translational outlook

We recently reported AAMs transplantation to improve the myocardium structurally and functionally after ischemic damage in a mouse model. ^11^ The multidisciplinary AADC consortium has successfully provided the leverage to translate these findings into clinical application initially as a clinical case report and description of methodology ^10^ and now here into the clinical safety and feasibility evaluation in support to CABG surgery. These results have thus paved the way for a more extensive further assessment of AAMs transplantation in terms of efficacy and safety, again relying on the wide cross-disciplinary expertise of AADC consortium partners.

## Data Availability

Data available on request due to privacy/ethical restrictions

## Acknowledgements

Research nurses Liisa Blubaum, Mariitta Salmi, Anna Blubaum and Merja Autti are gratefully acknowledged and thanked for their professional expertise in the development of the protocol. MSc Sole Lätti is acknowledged for the graphics illustrations. The authors thank Dr. Jennifer Rowland for linguistic editing of the manuscript.

## Declaration of Helsinki

Authors state that the study complies with the Declaration of Helsinki. The trial protocol has been evaluated and approved by the Surgical Ethics Committee of the Hospital District of Helsinki and Uusimaa (number 180/13/03/02/13).

## Abbreviations

AB-BE: Arterial astrup base excess
AAMs: Autologous atrial appendage micrografts
ARB: Angiotensin receptor blocker
BMI: Body Mass Index
CABG: Coronary artery bypass grafting
CCB: Calcium channel blockers
CMR: Cardiac magnetic resonance imaging
COPD: Chronic Obstructive Pulmonary Disease
CVP: Central venous pressure
ECG: Electrocardiogram
ECM: Extracellular matrix
EDV: End diastolic volume
ESV: End systolic volume
EF: Ejection fraction
FWHM: Full-width at half-maximum
GFR: Glomerular filtration rate
Hb: Hemoglobin
LA: Left atrium
LBBB: Left bundle branch block
LV: Left ventricle
LVEDD: Left ventricular end diastolic diameter
MAP: Mean arterial pressure
MRA: Mineralocorticoid receptor antagonists
NYHA: New York Heart Association
NT-PRO-BNP: N-terminal pro-B-type natriuretic peptid
PCI: Percutaneous coronary intervention.
SD: Standard deviation above the mean
SI: signal intensity
SPO2: Peripheral capillary oxygen saturation
STRM: signal threshold versus reference mean

## References

1. Hillis LD, Smith PK, Anderson JL, et al. 2011 ACCF/AHA Guideline for Coronary Artery Bypass Graft Surgery: a report of the American College of Cardiology Foundation/American Heart Association Task Force on Practice Guidelines. Circulation 2011;6:652–735.

2. Hueb W, Lopes N, Gersh BJ, et al. Ten-year follow-up survival of the Medicine, Angioplasty, or Surgery Study (MASS II): a randomized controlled clinical trial of 3 therapeutic strategies for multivessel coronary artery disease. Circulation 2010;122:949–957.

3. Fisher SA, Doree C, Mathur A, Martin-Rendon E. Meta-analysis of cell therapy trials for patients with heart failure. Circ Res 2015;116:1361–77.

4. Nguyen PK, Rhee J, Wu JC. Adult Stem Cell Therapy and Heart Failure, 2000 to 2016: A Systematic Review. JAMA Cardiol 2016;1:831–841.

5. Cheng K, Wu F, Cao F. Intramyocardial autologous cell engraftment in patients with ischaemic heart failure: a meta-analysis of randomised controlled trials. Heart Lung Circ 2013;22:887–894.

6. D’Alessandro DA, Michler RE. Current and future status of stem cell therapy in heart failure. Curr Treat Options Cardiovasc Med 2010;12:614–627.

7. Radisic M, Christman KL. Materials science and tissue engineering: repairing the heart. Mayo Clin Proc 2013;88:884–898.

8. Stevens KR, Kreutziger KL, Dupras SK, et al. Physiological function and transplantation of scaffold-free and vascularized human cardiac muscle tissue. Proc Natl Acad Sci USA 2009;106:16568–16573.

9. Oberwallner B, Brodarac A, Anić P, et al. Human cardiac extracellular matrix supports myocardial lineage commitment of pluripotent stem cells. Eur J Cardiothorac Surg 2015;47:416–425.

10. Lampinen M, Nummi A, Nieminen T, Harjula A, Kankuri E. Intraoperative processing and epicardial transplantation of autologous atrial tissue for cardiac repair. J Heart Lung Transplant 2017;36:1020–1022.

11. Xie Y, Lampinen M, Takala J, et al. Epicardial transplantation of atrial appendage micrograft patch salvages myocardium after infarction. J Heart Lung Transplant 2020;39:707–718.

12. Nummi A, Nieminen T, Pätilä T, et al. Epicardial delivery of autologous atrial appendage micrografts during coronary artery bypass surgery-safety and feasibility study. Pilot Feasibility Stud 2017;3:74.

13. Pätilä T, Lehtinen M, Vento A, et al. Autologous bone marrow mononuclear cell transplantation in ischemic heart failure: a prospective, controlled, randomized, double-blind study of cell transplantation combined with coronary bypass. J Heart Lung Transplant 2014:3;567–574.

14. Lehtinen M, Pätilä T, Vento A, et al. Prospective, randomized, double-blinded trial of bone marrow cell transplantation combined with coronary surgery - perioperative safety study. Interact Cardiovasc Thorac Surg 2014:19;990–996.

15. Sintonen H. The 15D instrument of health-related quality of life:properties and applications. Ann Med 2001;33:328–336.

16. Vermes E, Childs H, Carbone I, Barckow P, Friedrich MG. Auto-threshold quantification of late gadolinium enhancement in patients with acute heart disease. J Magn Reson Imaging 2013;37:382–390.

17. Zhang L, Huttin O, Marie PY, et al. Myocardial infarct sizing by late gadolinium-enhanced MRI: Comparison of manual, full-width at half-maximum, and n-standard deviation methods. J Magn Reson Imaging 2016;44:1206–1217.

